# Syrona: Visual Analytics for Systematic Comparison of Health Datasets on the OMOP Common Data Model

**DOI:** 10.64898/2026.05.04.26352350

**Authors:** Maarja Pajusalu, Marek Oja, Kerli Mooses, Silver Heinsar, Triin Laisk, Taavi Tillmann, Sven Laur, Sulev Reisberg, Jaak Vilo, Raivo Kolde

**Affiliations:** Institute of Computer Science, University of Tartu, Estonia; STACC, Estonia; Heart Clinic, Institute of Clinical Medicine, University of Tartu; Heart Centre, Internal Medicine Clinic, North Estonia Medical Centre; and Critical Care Research Group, Institute of Molecular Bioscience, The University of Queensland; Estonian Genome Centre, Institute of Genomics, University of Tartu, Estonia; Institute of Family Medicine and Public Health, University of Tartu, Estonia

**Keywords:** OMOP CDM, SNOMED CT, Data Analysis, Reasoning, and Decision Making, Comparison, Coordinated Views

## Abstract

Health research increasingly relies on observational datasets that capture systematically different patient populations, affecting the representativeness, comparability, and transportability of study findings. Yet these differences are rarely quantified across clinical domains or explored interactively, and the workflow for pairwise dataset comparison remains fragmented. We present Syrona, a visual analytics workflow for systematic pairwise comparison of datasets and subcohorts on the OMOP Common Data Model. Syrona extracts annual prevalence for conditions, procedures, and drugs from any OMOP CDM database, computes prevalence ratios across demographic strata, and synthesizes them via multilevel meta-analysis. A coordinated dashboard - distributional overviews, stratified heatmaps, forest plots, and absolute prevalence comparisons - enables interactive exploration driven by domain-adaptive SNOMED CT and ATC filtering. Three case studies on Estonian national health data (495,000 persons, 2012-2024) demonstrate how the workflow supports representativeness assessment, institutional practice comparison, and coding artifact detection. Syrona is open-source and applicable to any OMOP CDM database. Explore the demo at: http://omop-apps.cloud.ut.ee/ShinyApps/Syrona/

## 1 Introduction

Health research increasingly depends on observational data from diverse sources: national health registries, biobanks, local care-site databases. Each source captures a different slice of the population, shaped by recruitment strategy, coverage criteria, and data collection methods. When researchers use these datasets interchangeably - or draw conclusions from one that are assumed to generalize to others - implicit differences in cohort composition can introduce bias. Yet these differences are rarely quantified systematically.

The OMOP Common Data Model (CDM) enables distributed research within the Observational Health Data Sciences and Informatics (OHDSI) community, spanning more than 330 data sources with 2.1 billion patient records across 34 countries [19]. The OHDSI ecosystem provides tools for single-database characterization and quality assessment, but its emphasis remains on profiling one database at a time. To our knowledge, no existing OHDSI tool provides an integrated visual analytics workflow for exploratory pairwise comparison of concept prevalence patterns between datasets (Section 2.1).

The comparison of two datasets stratified into demographic and temporal groups produces an overwhelming volume of results - thousands of clinical concepts across demographic strata - that cannot be interpreted without statistical synthesis. We previously validated the prevalence-ratio approach on Estonian data using the International Classification of Diseases version 10 (ICD-10) diagnosis codes, custom SQL, and R Shiny dashboard [15], confirming clinically meaningful results - but the implementation was non-portable and locked to one coding system.

Syrona generalizes this into a portable, multi-domain visual analytics system for any OMOP CDM database, covering conditions, procedures, and drugs with domain-specific vocabulary enrichment. Our contributions are:

1. **A portable visual analytics workflow** for systematic pairwise comparison of OMOP CDM health datasets and subcohorts, spanning aggregated extraction, multi-level statistical synthesis, and coordinated interactive views for overview, stratified comparison, and temporal inspection across conditions, procedures, and drugs.
2. **Domain-adaptive filtering through SNOMED CT compositional attributes** - leveraging anatomical site, morphology, device, method, and Anatomical Therapeutic Chemical (ATC) classification [28] to enable discovery paths along dimensions unavailable in flat classification systems, tailored per clinical domain.
3. **Decision-support utility in realistic comparison tasks**. Using national-scale Estonian health data, we demonstrate that the workflow supports three decision-relevant analytical tasks: identifying cross-dataset selection effects, distinguishing institutional coding or practice variation from plausible burden differences, and detecting coding transitions over time.

## 2 Related Work

### 2.1 OHDSI Ecosystem and Comparative Database Analysis

The OHDSI community maintains the OMOP Common Data Model (CDM) [18], a standardized framework where local source codes map to standard concepts from clinical terminologies including SNOMED CT for clinical findings and procedures [22], RxNorm (medications) [13], and others. OHDSI provides single-database tools: ACHILLES [8] computes aggregate characterization statistics (e.g., concept counts by year, age, sex), ATLAS [5] provides a web interface for cohort definition and study design, and the Data Quality Dashboard (DQD) [3] performs automated conformance and completeness checks.

In addition, several open-source R packages have been developed to accelerate the use of OMOP CDM databases in health research and support the visualisation of results. However, there is a lack of tools that enable the comparison of several datasets. There are some packages used in OHDSI network studies which compare databases for specific clinical questions [9, 23]. CohortDiagnostics [17] is the closest tool, supporting multidatabase cohort evaluation, but its focus is on phenotype diagnostics in visualized tabular format - it does not compute any difference measures like prevalence ratios with meta-analytic pooling or support vocabulary-guided exploration of thousands of concepts. To our knowledge, no existing OHDSI tool provides an integrated visual analytics workflow for exploratory pairwise comparison of concept prevalence patterns between datasets.

### 2.2 SNOMED CT and Vocabulary-Driven Analytical Exploration

SNOMED CT serves as the standard vocabulary for conditions and procedures in OHDSI [22]. Unlike ICD-10 [27], it supports poly-hierarchy and compositional attributes (anatomical site, morphology, method, device) stored as defining relationships in OMOP vocabulary tables [14]. Hao et al. [7] showed that hierarchy quality directly impacts cohort query recall and precision. SNOMED has been used in OMOP CDM as a structure for standardization in clinical coding, cohort definition, and database characterization. While ICD-10 provides a single axis, SNOMED attributes enable multi-axis queries (e.g., “all procedures affecting heart structure”). Syrona builds on this property by using vocabulary structure as an analytical navigation layer for comparative exploration.

### 2.3 Visual Analytics for Health Data

Recent studies confirm that most health visualization tools provide descriptive summaries rather than comparative analytics with statistical inference. Wang and Laramee [25] reviewed 99 EHR visualization systems in depth, finding that most focus on single-patient or single-database views. Chishtie et al. [4] synthesized 56 population health visualization applications, observing limited integration of analytical inference into interactive exploration. Syrona adds to existing visualization solutions by combining interactive exploration and analytical interface for comparing two datasets.

Effective dashboard design relies on the foundational principle of “overview first, details on demand” [21]. In modern business intelligence dashboards, this is typically implemented through interactive multi-panel layouts that allow users to drill down from macro-level summaries into granular data [1, 12]. Syrona applies these established design patterns - from overview to detail [21], faceted displays [1, 10, 12], heatmaps [26], and forest plots [11]. To our knowledge, no existing tool combines multi-domain clinical comparison, pairwise statistical inference via meta-analysis, vocabulary-driven compositional filtering, and coordinated interactive visualization.

## 3 Problem Characterization and Design Requirements

### 3.1 Stakeholders and Analytical Tasks

Four analytical tasks, identified through collaboration with health data scientists and clinical specialists, motivated Syrona’s design:

- First, cross-dataset representativeness assessment: comparing prevalence patterns between two OMOP CDM datasets to assess differences that may influence comparability and transportability of study findings. This is a core concern for OHDSI network study facilitators.
- Second, selection bias assessment: comparing a representative population dataset against a voluntary biobank cohort to assess the direction and magnitude of participation-related selection effects.
- Third, care-site specialization assessment: comparing condition and procedure prevalence across hospital cohorts within the same dataset to quantify institutional practice variation.
- Fourth, coding practice change detection: inspecting per-year, age-stratified prevalence for related concepts to distinguish coding transitions from genuine clinical changes.

These tasks span different stakeholder roles but share a common analytical need: systematic, domain-aware pairwise comparison with statistical synthesis. They are demonstrated empirically in Section 5.

### 3.2 Design Requirements

The requirements for Syrona were derived from long-term collaboration with health data scientists and validated through iterative prototype testing, including a prior ICD-10-based pilot [15]. The development was motivated by a recurring bottleneck in federated network studies: the need to assess cross-dataset representativeness and data compatibility before investing in complex, multi-site cohort design.

In this context, domain experts must understand not just *if* datasets differ, but by how much, in which specific demographic strata, and whether these differences reflect clinical reality or data artifacts. Navigating thousands of concept-level comparisons across multiple clinical domains required a structured approach to managing visual and cognitive complexity. While the need for population-level overviews and granular demographic drill-downs drove the initial technical scope (**R1– R3**), observing clinician interactions with early prototypes revealed the necessity for visual clarity under scale and vocabulary transparency (**R4, R6**). Finally, the sensitive nature of real-world health data necessitated a privacy-preserving and portable architecture (**R7**). These observations were synthesized into the following seven design requirements:

#### R1. Multi-level comparison

Users must be able to navigate from population-level distributional overviews, through filtered concept groups, down to individual concepts stratified by sex, age group, and year, as concepts may show no overall difference but diverge significantly in specific strata.

#### R2. Statistical synthesis as visual simplification

Multi-level meta-analysis must reduce hundreds of stratum-level estimates into interpretable summaries. This acts as a dimensionality reduction step to enable scalable overview and heatmap views rather than a novel inferential contribution.

#### R3. Domain-adaptive semantic filtering

Filtering must extend beyond flat classification by leveraging SNOMED CT compositional attributes (anatomical site, morphology, device, method) and the ATC hierarchy, adapting dynamically per clinical domain.

#### R4. Visual clarity under scale

To manage thousands of simultaneous comparisons, the system must utilize progressive disclosure, restrained encoding (maximizing the data-ink ratio), and a consistent design language across all views.

#### R5. Flexible comparison scope

The workflow must support both cross-dataset comparisons (e.g., population sample vs. biobank) and within-dataset comparisons (e.g., hospital cohorts) without modification to the analytical pipeline.

#### R6. Concept search and vocabulary transparency

Users must be able to find concepts by name or code and inspect underlying vocabulary mappings to distinguish genuine clinical signals from administrative coding artifacts.

#### R7. Privacy-preserving portability

To ensure cross-site usability, only aggregated counts may leave the local database environment. A *k*-anonymity step (*k* = 5) must suppress sparse cells, and the pipeline must remain agnostic to specific SQL dialects across different OMOP CDM installations.

## 4 Syrona: Analytical Workflow and System Design

Syrona implements a three-phase pipeline (Fig. 2) source data extraction from any OMOP CDM database, 2) prevalence ratio computation between paired datasets, and 3) multi-level meta-analysis to synthesize stratum-level estimates into summary effects. A coordinated dashboard then enables interactive exploration of the results across three clinical domains.

**Fig. 1:**
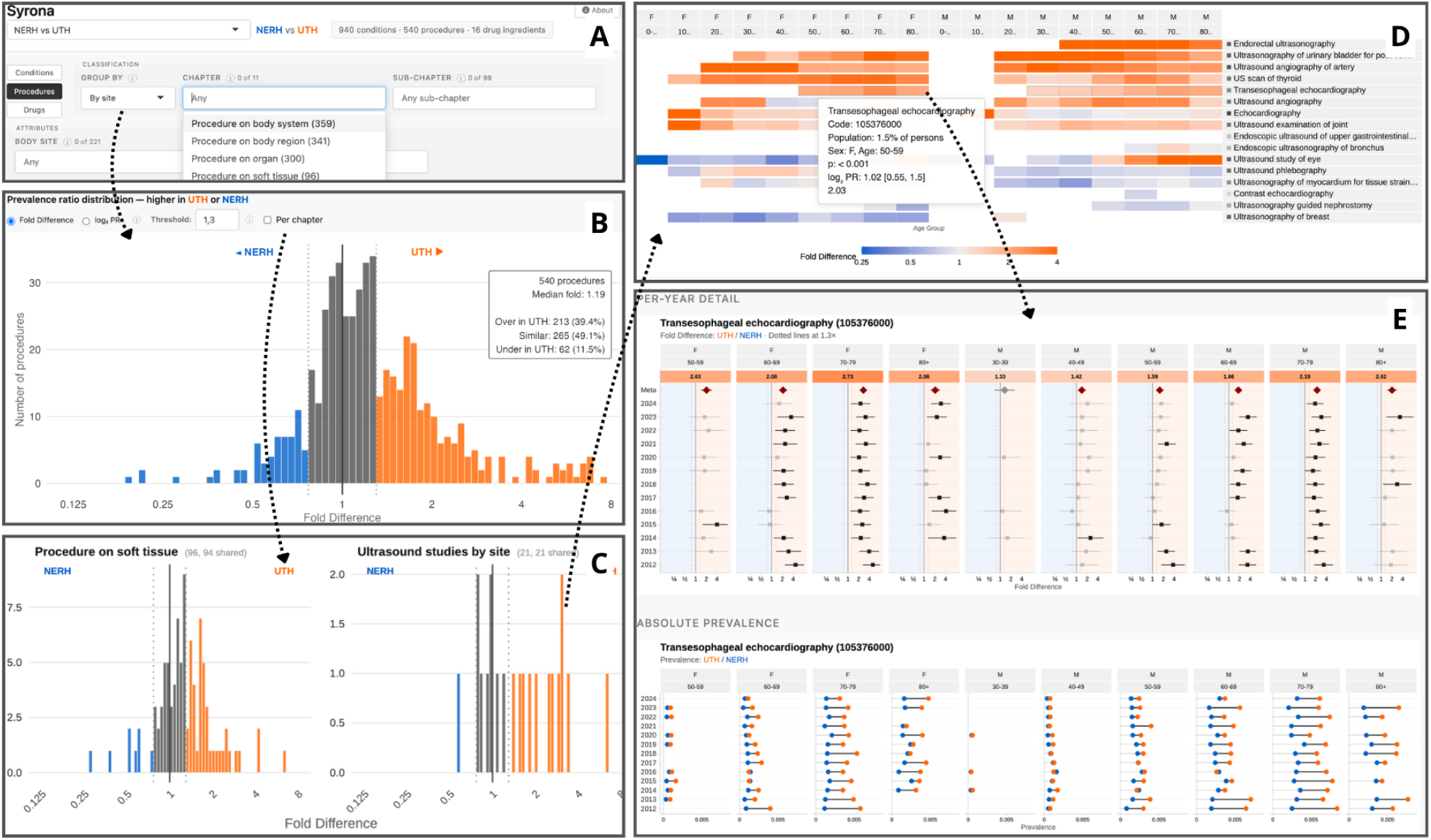
Syrona supports progressive exploration of prevalence differences between OMOP CDM health datasets, here comparing two Estonian hospitals (NERH and UTH). (A) The dashboard interface with domain and classification filters. (B) An overall prevalence ratio distribution reveals systematic differences across 508 procedures - blue indicates higher prevalence in NERH, orange in UTH. (C) Chapter-level faceted distributions isolate differences by clinical grouping. (D) A heatmap ranks individual procedures by fold difference; hovering reveals summary statistics and links to detail. (E) A stratified forest plot for a selected procedure (transesophageal echocardiography) shows prevalence ratios by sex and age group with meta-analytic pooling, alongside absolute prevalence comparison. The workflow guides analysts from population-level distributional patterns to concept-level stratified evidence.

**Fig. 2:**
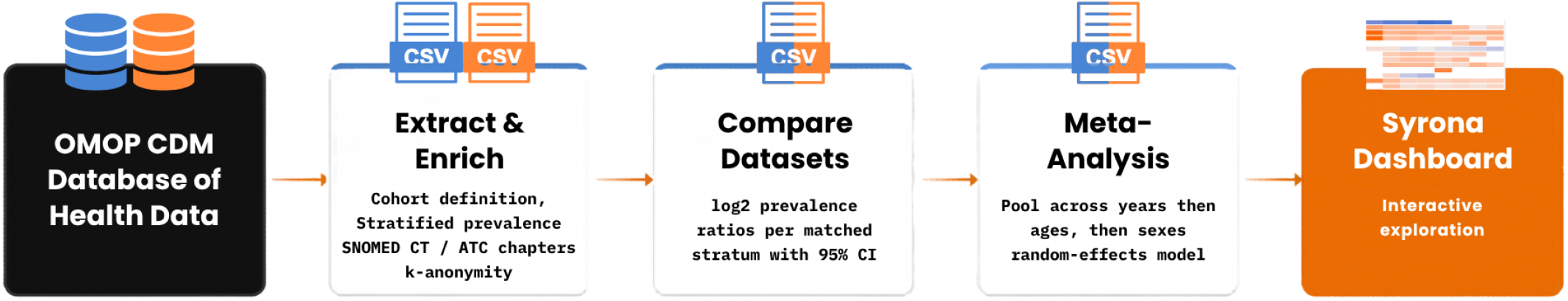
The Syrona computational architecture and data flow, moving from the OMOP CDM database through extraction, statistical synthesis, and final visual rendering.

### 4.1 Data Extraction and Comparison Setup

For each domain, Syrona extracts data from any OMOP CDM database using CDMConnector [2] and dplyr for portability : a prevalence fact table (one row per concept x year x sex x age group, with patient count and denominator) (R7), plus dimension tables for concept metadata, chapter classifications, and compositional attributes (**R6**). All three domains (conditions, procedures and druga) share the same fact table structure but differ in vocabulary axes: conditions use SNOMED CT with ICD-10 chapter and body system groupings; procedures use SNOMED CT with method- and site-based groupings; drugs use RxNorm ingredients mapped to ATC classification. A *k*-anonymity step (*k* = 5) suppresses cells with fewer than 5 patients (**R7**). The denominator for each stratum is derived from the OMOP observation_period table: a person contributes to a year if their observation period overlaps that calendar year; for cohort-based extractions, observation periods are clipped to the cohort window. Sub-cohorts (e.g., by care site) use OHDSI-standard cohort tables (**R5**).

### 4.2 Annual Prevalence and Prevalence Ratios

The underlying metric is the annual prevalence. For each concept, the proportion of persons in a stratum (age group x sex x calendar year) with at least one recorded occurrence during that year, is calculated. This definition applies uniformly across conditions, procedures, and drug ingredients, placing all three domains on a comparable person-based scale (**R2**). It captures the *reach* of a clinical concept rather than the *intensity* of service use. Repeated events, dosage, and utilization volume are intentionally not represented. This trade-off is further discussed in Section 7.2. This person-level perspective aligns with OHDSI characterization practice [8].

For each matched stratum, Syrona computes a log_2_ prevalence ratio with 95% confidence interval. The log_2_ scale is symmetric (2x maps to +1, 0.5x to -1, equal prevalence to 0); back-transformation yields the fold difference shown in the dashboard. A default fold threshold of 1.3, calibrated empirically based on previous analysis, defines the “similar” zone. The threshold is a visual reference, adjustable interactively.

### 4.3 Multilevel Aggregation and Summaries

Per-stratum log_2_ prevalence ratios are pooled in three sequential levels via random-effects meta-analysis (Paule-Mandel estimator for between-study variance [16], implemented via the R *meta* package [20]): first across years within each age-sex group, then across age groups within each sex, then across sexes - yielding one summary estimate per concept. At each level, the pipeline computes heterogeneity metrics (*I*^2^, *τ*^2^, Q statistic) to quantify between-stratum variation.

This pooling serves primarily as visual simplification (R2): reducing hundreds of stratum-level estimates into navigable summaries that make the overview and heatmap views possible at scale, while preserving intermediate results for drill-down (**R1**). Because yearly estimates from overlapping populations are not fully independent, pooled summaries are used as descriptive aggregations for navigation rather than formal inferential tests, with per-year estimates remaining the primary analytical output. A fallback chain (random-effects→ fixed-effect → pass-through) ensures coverage under sparse data.

### 4.4 Vocabulary-Driven Filtering

Flat classification systems such as ICD-10 provide a single grouping axis. SNOMED CT’s compositional structure offers multiple orthogonal axes stored as defining relationships in OMOP vocabulary tables (**R3**). Syrona extracts these relationships at the concept level during Phase 1 and exposes them as cascading filter dimensions in the dashboard.

For conditions, three relationship types become filters: finding site (anatomical location), associated morphology (pathological process), and clinical course (acute, chronic, progressive). For procedures, four filter types are available: direct procedure site, method, device, and morphology. Drug ingredients can be filtered by the Anatomical Therapeutic Chemical (ATC) classification [28] with 14 chapters (i.e sensory organs, nervous system, blood and blood forming organs).

Filters cascade: selecting a chapter narrows available sub-chapters; selecting sub-chapters narrows attribute choices. This enables multi-axis queries unavailable in flat systems - for example, filtering cardiovascular procedures by device type to isolate pacemaker-related activity, or filtering conditions by finding sites to compare respiratory diagnoses across hospitals. Each filter combination propagates through all downstream views (**R1, R4**).

### 4.5 Dashboard Design and Coordinated Interactions

The dashboard implements coordinated views (**R1, R4**) linked by shared selections and driven by cascading filters (**R3**), following an overview-to-detail workflow. The interface is organized into four main components:

#### Global Controls

A domain selector switches between conditions, procedures, and drugs, adapting all filters and views (**R3**). Users can utilize the concept search for direct lookup by name or code (**R6**) or switch comparison datasets via a header selector (**R5**).

#### Macro-level Distributional Overview

The distribution of prevalence ratios provides the first entry point: a histogram of pooled fold differences, colored directionally (blue = reference-higher, orange = comparison-higher) with significance encoded by opacity. A “per chapter” toggle facets the distribution by clinical chapter; clicking a chapter automatically navigates to the pre-filtered heatmap.

#### Mid-level Stratified Heatmap

The heatmap shows one row per concept with columns faceted by sex x age group, encoding log_2_ PR on a diverging blue-gray-orange scale. Interactive ranking buttons allow users to surface the most over- or underrepresented concepts to prioritize exploration (**R4**).

#### Micro-level Detail View

Selecting a concept opens a temporal detail pane (Fig. 3). The top panel provides a forest plot showing per-year prevalence ratio estimates with 95% CIs and meta-analysis diamonds, providing the temporal resolution required for coding transition detection. Below, a prevalence dumbbell chart displays absolute prevalence values, revealing whether large fold differences reflect meaningful rates or amplified low-base-rate ratios.

**Fig. 3:**
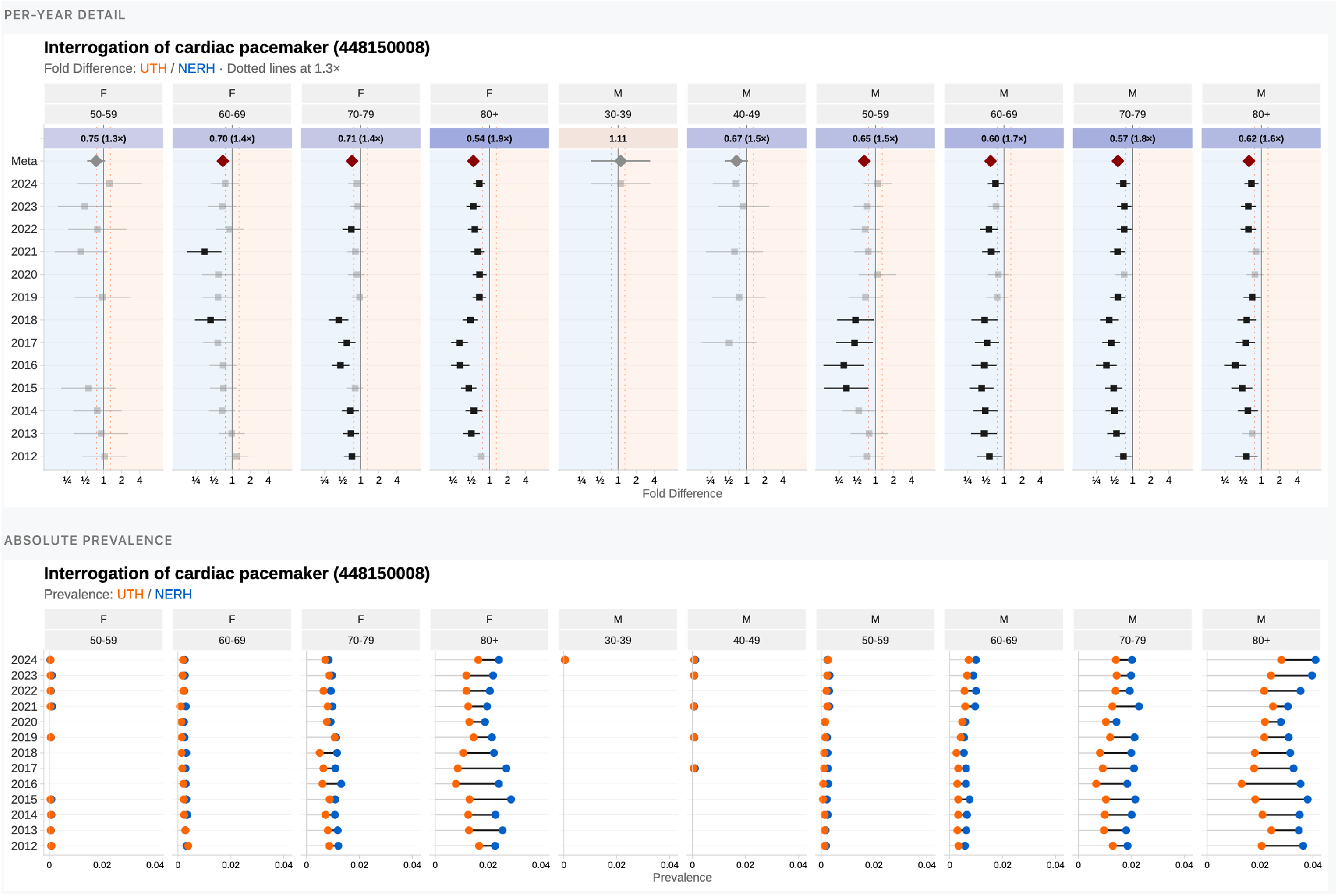
Detail-on-demand for “Interrogation of cardiac pacemaker” (SNOMED 448150008). Top: per-year log_2_ PR with 95% CI, faceted by sex x age group. Bottom: absolute prevalence dumbbell chart.

#### Design Evolution of Detail Views

Early iterations of the prototype utilized grouped bar charts to compare prevalence across datasets. However, experts found the high visual density of bars—stratified by sex, age, and year—to be cluttered and prone to occlusion (**R4**). We transitioned to the *dumbbell chart* encoding to minimize visual weight while preserving the directional relationship between datasets. By replacing thick bars with a light connector line and color-coded dataset markers, we maximized the data-ink ratio. This encoding allows users to quickly identify temporal patterns through the geometric arrangement of the markers: vertical alignments indicate stability, diagonal shifts suggest gradual prevalence changes, and “snake-like” patterns reveal fluctuating clinical or coding trends over time.

The dashboard is implemented in R Shiny with ggiraph [6] for interactive SVG graphics. A consistent color scheme (blue = reference, orange = comparison) is maintained across all views to reduce cognitive load.

## 5 Case Studies

We demonstrate Syrona on Estonian health data using two comparison settings: cross-dataset and within-dataset. Est-Health-30 is a 30% random sample of the Estonian population from national health insurance claims and prescriptions. The Estonian Biobank (EstBB) includes volunteer participants linked to the same claims and prescriptions data, reflecting self-selection into biobank recruitment. Hospital cohorts are derived from Est-Health-30 using OHDSI-standard cohort definitions, selecting individuals with at least one visit to a given hospital; clinical events are restricted to those recorded at that hospital (care-site filtering), isolating institutional practice from care received elsewhere.

### 5.1 Cross-Dataset Comparison: Representative Population vs. Biobank

Comparing Est-Health-30 with the EstBB reveals a selection effect consistent across all three domains (Fig. 4): most concepts are over-represented in EstBB (median fold 1.32-1.35), while fewer than 5% are underrepresented. Overrepresented concepts are consistent with a health-engaged population. Underrepresented concepts cluster around addiction, severe mental illness, and intellectual disability. Switching domains requires a single button click; the consistent shift across 4,664 concepts in three independent clinical vocabularies becomes visible within seconds - a pattern that would require three separate analyses and manual comparison in a tabular workflow.

**Fig. 4:**
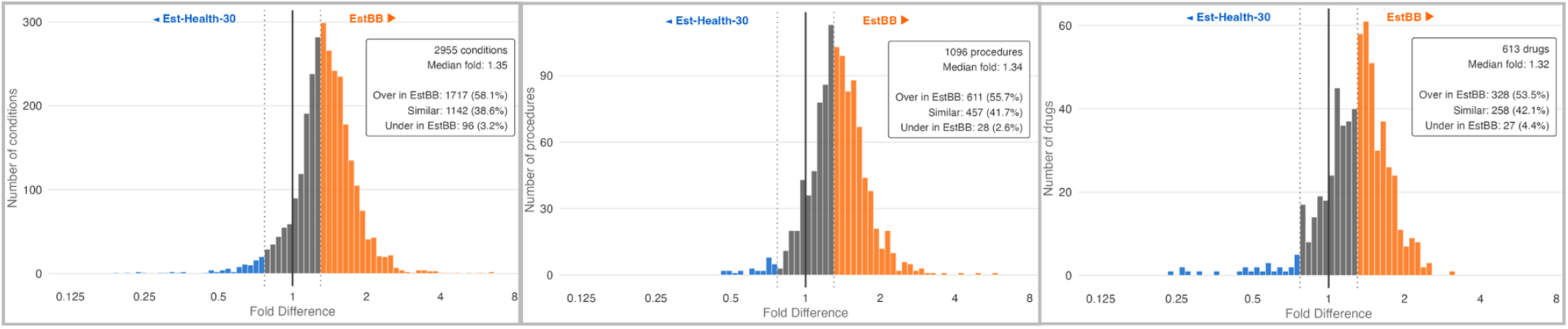
Cross-dataset comparison: Est-Health-30 (495,000 persons) vs. Estonian Biobank (212,000 volunteers). PR distributions for conditions (A), procedures (B), and drugs (C). Rightward shift indicates higher prevalence in EstBB.

For a network study facilitator assessing whether these datasets can be used interchangeably in a distributed study, the distributional overview provides an immediate answer: the systematic rightward shift means EstBB overrepresents health-engaged individuals, and any cross-dataset analysis must account for this structural difference. For a biobank researcher, the same comparison quantifies the direction and magnitude of the healthy-volunteer effect - the multi-domain concordance confirms a structured selection bias reflected simultaneously in diagnoses, procedures, and medications, rather than driven by domain-specific coding effects.

### 5.2 Within-Dataset Comparison: Hospital Cohorts

This within-dataset comparison uses hospital-based subcohorts presenting two Estonian higher-level hospitals: North-Estonian Regional Hospital (NERH) situated in the capital and covering mostly the north and west part of the country, and University of Tartu Hospital (UTH) situated in the southern part of the country and covering mostly this region. Using Syrona workflow and cohort definition we extracted the data from Est-Health-30, eliminating cross-dataset selection bias since all cohorts originate from the same population sample. Care-site filtering then restricts events to those recorded at each hospital, isolating institutional coding and practice patterns. At the overview level, conditions diverge more than procedures: of 940 compared conditions, 45.8% are overrepresented at UTH, whereas procedures are more balanced, with 52.8% of 540 concepts falling within the similarity zone (Fig. 5A-B). This asymmetry suggests that institutional variation is stronger in diagnostic labeling than in procedural activity.

**Fig. 5:**
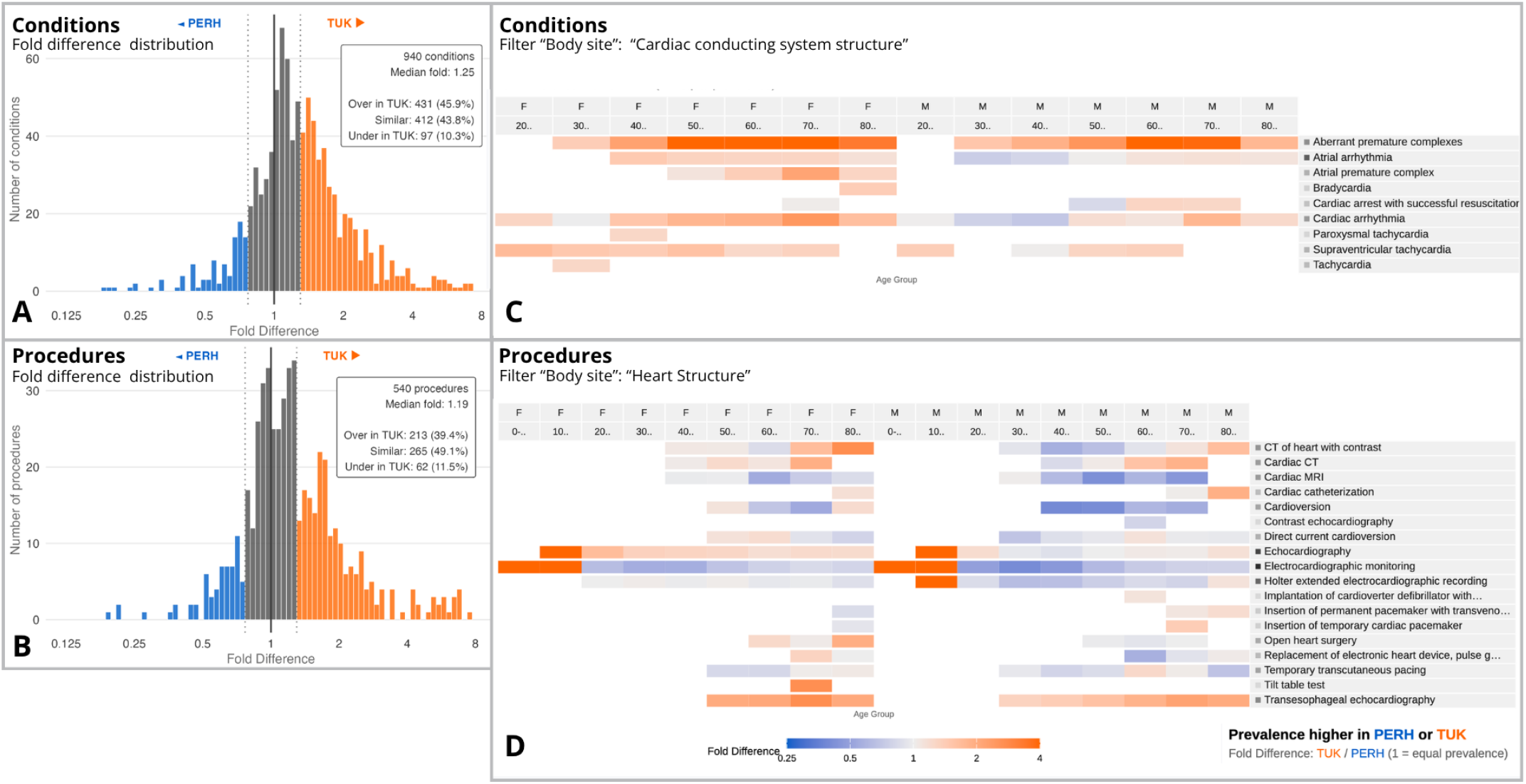
Within-dataset comparison: NERH vs UTH (care-site filtered). (A-B) PR distributions for 940 conditions and 540 procedures show stronger divergence in conditions than in procedures. (C-D) Drill-down via SNOMED CT attribute filtering.

Drilling into an example of cardiac rhythm disorders. Filtering conditions by SNOMED CT finding site to cardiac structures surfaces a pattern that illustrates this asymmetry at the concept level. Table 1 shows three cardiac rhythm conditions ordered by diagnostic specificity. Atrial arrhythmia (I48), which has precise electrocardiographic diagnostic criteria, is near parity across the two hospitals (fold 1.13, *p* = 0.286). Cardiac arrhythmia (I49) - a residual category that ICD-10 defines as “Other cardiac arrhythmias” - trends UTH-higher (fold 1.34, *p* = 0.142). Angina pectoris (I20), a broader clinical category, diverges further (fold 1.73, *p* = 0.015). This ordering is consistent with a specificity gradient: diagnostically precise codes converge across institutions, while broader residual codes absorb more of the institutional variation in coding practice.

**Table 1:** Cardiac rhythm conditions: NERH vs. UTH (care-site filtered). Fold = UTH/NERH prevalence ratio, pooled across age-sex strata via random-effects meta-analysis.

| Condition | SNOMED | ICD-10 | Fold | p-value |
| --- | --- | --- | --- | --- |
| Atrial arrhythmia | 17366009 | I48 | 1.13 | 0.286 |
| Cardiac arrhythmia | 698247007 | I49 | 1.34 | 0.142 |
| Angina pectoris | 194828000 | I20 | 1.73 | 0.015 |

The “specificity-gradient” in the cardiovascular data initially prompted specialist skepticism. The observed diagnostic discordance was surprising, particularly as non-specific residual codes exhibited lower prevalence than more granular, concordant codes. This inverted relationship suggested a localized coding bias rather than a general population difference. To resolve this, we conducted a cross-domain validation using procedural data. We hypothesized that if the diagnostic excess reflected a genuine clinical burden, one would expect a corresponding overrepresentation in rhythm-related therapeutic procedures.

Switching to the procedure domain and filtering by cardiac site reveals a different picture (Table 2). Core rhythm-oriented procedures - electrocardiographic monitoring (fold 1.01), Holter recording (0.96), and DC cardioversion (0.97) - are concordant across hospitals; general cardioversion is NERH-leaning (fold 0.55, *p <* 0.001). None of these procedures mirror the UTH diagnostic excess. Echocardiographic procedures tell a separate story: standard echocardiography (fold 1.61, *p* = 0.035) and transesophageal echocardiography (fold 2.02, *p* < 0.001) are both UTH-higher, reflecting institutional diagnostic workup preferences rather than rhythm disorder burden.

**Table 2:** Cardiac rhythm-related procedures: NERH vs. UTH (care-site filtered). Fold = UTH/NERH prevalence ratio, pooled across age-sex strata via random-effects meta-analysis.

| Procedure | SNOMED | Fold | p-value |
| --- | --- | --- | --- |
| Electrocardiographic monitoring | 46825001 | 1.01 | 0.969 |
| Holter recording | 427047002 | 0.96 | 0.153 |
| Cardioversion | 250980009 | 0.55 | <0.001 |
| DC cardioversion | 180325003 | 0.97 | 0.559 |
| Echocardiography | 40701008 | 1.61 | 0.035 |
| Transesophageal echocardiography | 105376000 | 2.02 | <0.001 |

The diagnosis-procedure dissociation confirms that the condition-level pattern alone could indicate either a true burden difference or a coding artifact, but the absence of a matching procedural excess points toward coding allocation as the dominant driver. This reasoning required switching between two clinical domains - conditions and procedures - within the same comparison, each filtered through SNOMED CT attributes to the relevant concept subset.

This case illustrates how Syrona supports within-dataset reasoning beyond simple hospital profiling. By linking condition- and procedure-level views through vocabulary-driven drill-down, the system helps distinguish case-mix differences from structured institutional coding variation - a conclusion that neither the condition-level pattern nor the procedure-level evidence would support alone (R1, R3, R6).

### 5.3 Detection of Coding Practice Change: Mammography

Syrona extracts two distinct SNOMED concepts: general mammography (71651007) and screening mammography (24623002). Comparing NERH and UTH reveals a coding practice transition visible only through per-year temporal resolution (Fig. 6).

**Fig. 6:**
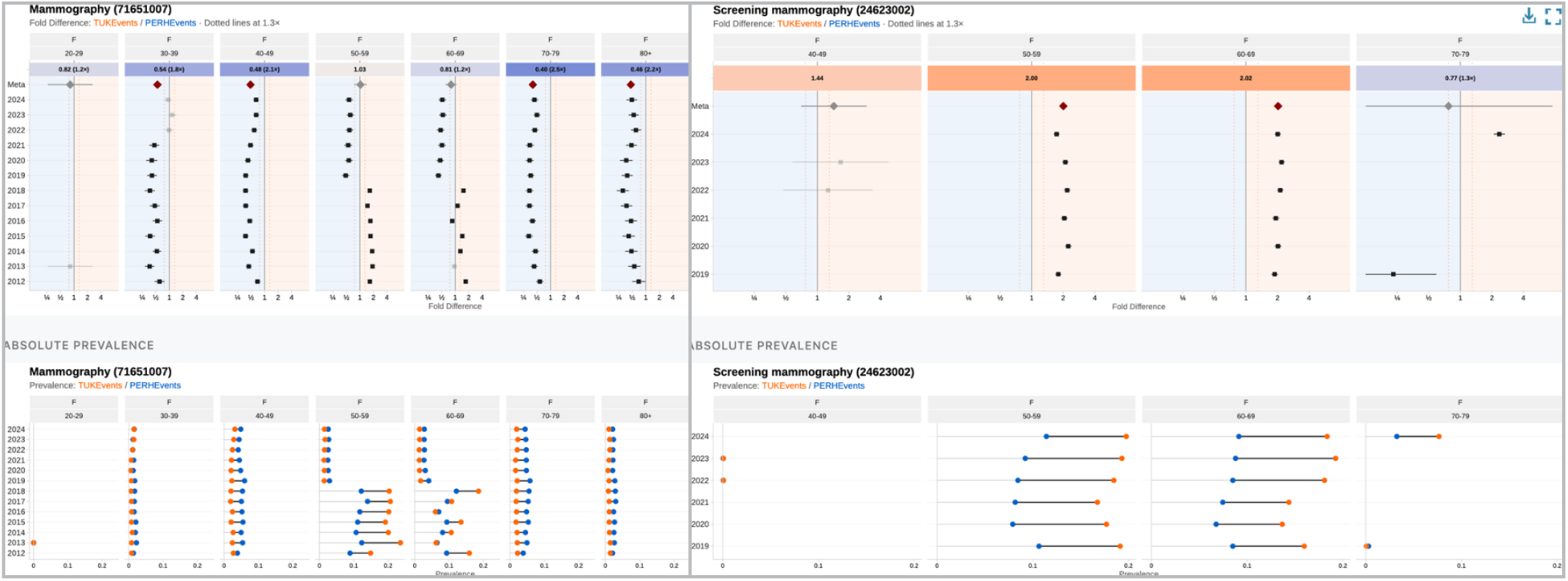
Coding practice detection: mammography (NERH vs. UTH, care-site filtered). Top: forest plots showing the post-2019 code transition. Bottom: absolute prevalence dumbbells. UTH leads in screening-age groups both before and after the transition.

Screening mammography shows zero records before 2019, then appears in the 50-69 screening target range (Fig. 6, top-right); general mammography shows a corresponding step-down in the same strata (Fig. 6, top-left). This confirms a code substitution, not a clinical change: screening was recorded under the general code until 2019, when a distinct screening code was adopted. Younger age groups are absent from the screening panel, consistent with the national program targeting women 50-69.

Beyond detecting the transition, the comparison reveals a substantive finding. UTH shows consistently higher screening prevalence in the target age range - and this lead is already visible before 2019 in the general mammography forest plot, when screening was still coded under the general concept (Fig. 6, top-left). The pattern persists across the coding transition, ruling out a coding-only explanation and indicating a genuine regional difference in screening uptake between NERH and UTH.

Without temporal stratification across related concepts, a researcher querying only “mammography” would observe a post-2019 decline and misinterpret it as reduced screening (**R1, R6**).

## 6 Expert Evaluation and Design Iterations

To assess the analytical efficacy of Syrona and refine its visual encodings, we conducted formative evaluation sessions with seven domain experts. Rather than a controlled laboratory experiment, we employed a think-aloud protocol and guided exploration to understand how experts interact with the system in realistic, open-ended analytical scenarios.

### 6.1 Participants and Methodology

Participants represented three distinct user archetypes within the OHDSI ecosystem:

- **Data Scientists (***n* = 3): Experts with 10+ years of experience in OMOP CDM, federated network studies, and data engineering in both Estonian and international settings.
- **Clinical Specialists (***n* = 4): Researchers specializing in cardiology, women’s health, general practitioner, and public health
- policy, representing the “report consumers” who initiate clinical research questions.
- **Biobank Researchers (***n* = 2): Specialists focused on population genetics and cohort representativeness.

Sessions were conducted as collaborative workshops. For the data scientists and biobankers, sessions involved hands-on, shared-screen interaction with the tool, ideating requirements, and stress-testing the interface with targeted queries. For the clinical specialists, sessions focused on interpreting generated visual reports to simulate their real-world role as hypothesis generators.

### 6.2 Sensemaking and Analytical Workflows

The evaluation revealed that different archetypes derived distinct forms of value from the system, validating its cross-disciplinary utility:

#### Organic Discovery of Artifacts

The data scientists immediately stress-tested the demographic stratifications. While exploring female health concepts, one engineer noted a sudden, suspicious drop in general mammography prevalence. By utilizing the search interface for the prefix “mammo…”, the “screening mammography” concept appeared. Seeing the exact inverse trend visually confirmed the administrative coding shift (detailed in Case Study 3).

#### Cross-Domain Hypothesis Generation

Clinical specialists validated the analytical loop. When presented with the unexplained diagnostic excess in cardiac arrhythmias (I49), they successfully used the tool to cross-reference therapeutic procedures, concluding the discrepancy was a coding artifact. Immediately following this, they asked, *“Can we see the blood tests?”* This confirmed the natural extension of the workflow and identified laboratory measurements as the most critical domain for future integration.

#### The Need for Macro-Level Context

While early iterations of the prototype provided detailed stratified differences and meta-analyzed concept views, feedback from the biobank researchers emphasized a missing layer of abstraction required to quantify the healthy volunteer effect.” Although they valued the granular clinical detail, their primary operational requirement was to understand the population-level bias: *How do we show the overall shift?”* This insight fundamentally validated the requirement for a macro-level distributional overview as the entry point for any comparative exploration (**R1**).

### 6.3 Iterative Design Refinements

Applying theoretical visualization guidelines to highly sparse, complex clinical data revealed several points of friction with expert mental models. These observations drove specific domain-adaptive design iterations (**R4**):

1. **Analytical Hierarchy (Overview First):** While early prototypes allowed immediate entry into granular heatmaps, biobank researchers prioritized quantifying overall cohort bias (e.g., the “healthy volunteer effect”). We enforced a strict Shneiderman mantra (overview first), requiring users to navigate the macro-level distributional histograms before accessing concept-level details.
2. **Encoding Trade-offs (**log_2_ **vs. Fold Difference):** Theoretically, log_2_ prevalence ratios provide optimal visual symmetry for encoding relative differences. However, clinical specialists struggled with its cognitive interpretability during rapid screening. We optimized for the clinical mental model by elevating absolute Fold Difference as the primary dashboard metric, retaining log_2_ encoding only for the underlying meta-analytical rendering.
3. **Addressing Sparse-Data Ambiguity in Color Scales:** Standard diverging color scales frequently utilize white as a neutral midpoint. However, in heavily stratified observational health data, sparsity is common. Users conflated white (parity) with background canvas (missing data). We deviated from standard palettes by anchoring the neutral midpoint to light gray, explicitly disambiguating ‘zero effect’ from ‘missing data’.
4. **Scaffolding Complex Ontologies:** Migrating from ICD-10 to SNOMED CT ensured international portability but introduced significant cognitive load due to poly-hierarchical relationships. Because standard cascading filters obscure these mappings, we implemented a persistent vocabulary provenance table, allowing users to verify hierarchy paths and trust the filtering output (**R6**).

## 7 Discussion

### 7.1 What Syrona Changes in Practice

The OHDSI ecosystem excels at answering pre-specified clinical questions across databases. Syrona addresses a significant need during the design stage of study: exploratory comparison, asking where datasets differ and why rather than estimating a specific clinical effect. This shifts analysis from hypothesis-driven cohort studies toward structural profiling that informs study design and interpretation. The same pipeline supports cross-dataset and within-dataset comparison across conditions, procedures, and drugs without modification, while sharing only aggregated counts outside the database environment. While the case study examples were produced using Estonian datasets, we also validated the full workflow on Synthea synthetic data [24], confirming that the pipeline produces the expected output schema from a non-Estonian OMOP CDM installation without any code modification.

### 7.2 What the Metric Choice Enables and Hides

Person-level prevalence asks *what proportion of people are affected or exposed*; event counts ask *how much service is delivered*. Both are valid epidemiological perspectives. Syrona uses person-level annual prevalence because its goal is comparative profiling - detecting which concepts are over- or underrepresented between populations – not measuring service volume. This choice standardizes conditions, procedures, and drugs onto one comparable scale, prevents high-frequency users from dominating the signal, and supports fairer comparison when datasets differ in recording density. It is well-matched to the case studies: biobank representativeness (Section 5.1) is inherently a person-level question, and the mammography coding transition (Section 5.3) tracks code adoption, not procedure counts.

The trade-off is real for highly recurrent services - dialysis, chemotherapy, repeat imaging - where intensity differences are clinically meaningful but invisible to person-level prevalence. For drugs, prevalence of exposure does not capture dose, adherence, or duration. These limitations are inherent to the metric, not to the system: Syrona extracts event counts alongside prevalence, and an event-rate mode is a natural next step (Section 7.4).

### 7.3 Limitations

Syrona currently supports pairwise comparison only; extending to network-wide matrices of multiple datasets would require rethinking the visual layout. The person-level prevalence metric does not capture repeated-event intensity (Section 7.2). Because yearly estimates from overlapping populations are not fully independent, pooled meta-analytic summaries serve as descriptive aggregations for navigation rather than formal inferential tests - per-year stratified estimates remain the primary analytical output.

The dashboard encodes statistical significance through opacity but does not yet visualize heterogeneity. This limits users’ ability to distinguish stable effects from those where the pooled estimate masks between-stratum variation (see Section 7.4). Because of the SNOMED CT poly-hierarchy, a single concept may appear under multiple chapters. The current filtering treats these as inclusive unions, which can produce unexpected overlap in filtered views.

Results depend on the quality of source-to-standard concept mappings in each CDM installation. Incomplete or inconsistent mappings at the local site will propagate silently into the prevalence ratios. Finally, the expert evaluation (see Section 6) was conducted by the tool’s developer with a small number of domain experts. It does not constitute a formal user study.

### 7.4 Future Work

We identify three primary trajectories to extend the system’s utility and evaluative depth:

1. **Longitudinal Intensity Metrics:** While the current pipeline effectively captures event counts, we plan to integrate an event-rate mode. By visualizing events per 1,000 person-years, the system will move beyond person-level prevalence to address nuanced questions of utilization intensity (Section 7.2). This transition is vital for distinguishing between isolated occurrences and chronic clinical patterns.
2. **Visualizing Statistical Heterogeneity:** To improve the interpretability of aggregated data, we will expose existing measures of statistical heterogeneity through dedicated visual channels. Specifically, we intend to integrate prediction interval bands into the forest plots. These visual cues will allow users to immediately differentiate between stable, homogenous effects and those driven by underlying data heterogeneity.
3. **Expanded Expert Evaluation:** Beyond the initial case studies, we have identified additional clinical scenarios that warrant deeper investigation. We are currently transitioning into a broader evaluative phase to test these new system features with a more diverse cohort of specialists. This iterative approach will allow us to assess how the tool performs across distinct clinical workflows.

## 8 Conclusion

We presented Syrona, a visual analytics workflow for systematic pairwise comparison of OMOP CDM health datasets and subcohorts. Syrona integrates portable extraction, stratified prevalence ratios, multilevel meta-analysis, and vocabulary-driven filtering into a coordinated visual workflow. This allows domain experts to move from population-level distributional overviews to stratum-level and temporal detail across conditions, procedures, and drugs without modifying the analytical pipeline.

Three case studies on Estonian national health data demonstrated the analytical utility of the workflow across distinct comparison tasks: identifying selection effects, distinguishing coding allocation from plausible clinical differences, and detecting temporal coding transitions. Together, they show how Syrona combines statistical synthesis with interactive visual exploration to support scalable comparison, targeted drill-down, and more defensible interpretation.

Syrona is open source and applicable to any OMOP CDM database. By treating dataset comparison as a visual analytics problem rather than a tabular reporting task, it addresses a gap in the OHDSI ecosystem and supports the growing need for systematic assessment of datasets in distributed observational research. More broadly, Syrona helps experts reach more defensible conclusions about whether observed differences reflect selection effects, institutional practice variation, coding artifacts, or plausible clinical differences.

## Data Availability

All data produced are available online at https://github.com/MaarjaPajusalu/Syrona/

## Supplemental Materials

All supplemental materials, including source code and sample data, will be made available at https://github.com/MaarjaPajusalu/Syrona

## Figure Credits and Copyrights

All figures are original work by the authors.

## Data availability statement

The data analyzed in this study was obtained from the Estonian Biobank (EstBB). Individual level data from the Estonian Biobank or Est-Health-30 cannot be shared publicly due to legal and ethical restrictions. According to the Human Genes Research Act, which regulates the operations of the Estonian Biobank, the data include potentially identifiable genetic and health information and are therefore subject to restricted access. Access to these data is only possible directly through the Estonian Biobank upon ethics approval from the Estonian Committee on Bioethics and Human Research. Researchers may request access following the procedure described here: https://genomics.ut.ee/en/content/estonian-biobank#dataaccess and requests should be directed to. The data generated and used as aggregated data for the analysis can be accessed via the open-access diagnostic dashboard developed during this study https://github.com/MaarjaPajusalu/Syrona or contacting the authors directly.

## Ethics approval

This work was approved by the Estonian Bioethics and Human Research Council (1.1-12/653, 1.1-12/1039) and the Ethics Committee of University of Tartu (401/T-34).

## Acknowledgments

We wish to thank Gerhard Grents for sharing his expert insights, contributing to prototype testing and feedback.

## References

[1] B. Bach et al. Dashboard design patterns, 2023.

[2] A. Black, P. R. Rijnbeek, and E. Burn. Cdmconnector: An r package for working with observational health data in the omop common data model. Bioinformatics, 39(12):btad682, 2023. doi: 10.1093/bioinformatics/btad682

[3] C. Blacketer, F. J. Defalco, P. B. Ryan, and P. R. Rijnbeek. Increasing trust in real-world evidence through evaluation of observational data quality. Journal of the American Medical Informatics Association, 28(10):2251–2257, 2021. doi: 10.1093/jamia/ocab132

[4] J. Chishtie, I. A. Bielska, L. Engel, M. Beauséjour, S. Berthelot, T. Hartley et al. Interactive visualization applications in population health and health services research: Systematic scoping review. Journal of Medical Internet Research, 24(2):e27534, 2022. doi: 10.2196/27534

[5] F. J. DeFalco et al. Atlas: A unified interface for the ohdsi tools, 2022.

[6] D. Gohel and P. Skintzos. ggiraph: Make ‘ggplot2’ graphics interactive. r package version 0.8.9, 2024.

[7] B. Hao, Y. Li, M. Sotoodeh, and H. Yu. Quantitatively assessing the impact of the quality of snomed ct subtype hierarchy on cohort queries. Journal of the American Medical Informatics Association, 32(1):89–96, 2025. doi: 10.1093/jamia/ocae258

[8] G. Hripcsak, J. D. Duke, N. H. Shah, C. G. Reich, V. Huser, M. J. Schuemie et al. Observational health data sciences and informatics (ohdsi): Opportunities for observational researchers. Studies in Health Technology and Informatics, 216:574–578, 2015. doi: 10.3233/978-1-61499-564-7-574

[9] G. Hripcsak, P. B. Ryan, J. D. Duke, N. H. Shah, R. W. Park, V. Huser et al. Characterizing treatment pathways at scale using the ohdsi network. Proceedings of the National Academy of Sciences, 113(27):7329–7336, 2016. doi: 10.1073/pnas.1510502113

[10] W. Javed and N. Elmqvist. Exploring the design space of composite visualization. In Proc. IEEE Pacific Visualization Symposium, pp. 1–8, 2012. doi: 10.1109/PacificVis.2012.6183556

[11] S. Lewis and M. Clarke. Forest plots: Trying to see the wood and the trees. BMJ, 322(7300):1479–1480, 2001. doi: 10.1136/bmj.322.7300.1479

[12] Y. Lu et al. Interactive dashboards for visual analytics, 2021.

[13] S. J. Nelson et al. Rxnorm: A clinical drug vocabulary, 2011.

[14] A. Ostropolets, C. Reich, P. Ryan, and G. Hripcsak. Characterizing database granularity using snomed-ct hierarchy. In AMIA Annual Symposium Proceedings, pp. 983–992, 2021.

[15] M. Pajusalu et al. Comparison of the prevalence of all diagnosed diseases among estonian biobank participants against the general population, 2026. Preprint doi: 10.64898/2026.02.05.26345634.

[16] R. C. Paule and J. Mandel. Consensus values and weighting factors. Journal of Research of the National Bureau of Standards, 87(5):377–385, 1982. doi: 10.6028/jres.087.022

[17] G. Rao, M. J. Schuemie, P. B. Ryan, P. E. Stang, G. Hripcsak, and M. A. Suchard. Cohortdiagnostics: Phenotype evaluation across a network of observational data sources. PLoS ONE, 20(1):e0310634, 2025. doi: 10.1371/journal.pone.0310634

[18] C. Reich, A. Ostropolets, P. Ryan, P. Rijnbeek, M. Schuemie, A. Davydov et al. Ohdsi standardized vocabularies-a large-scale centralized reference ontology for international data harmonization. Journal of the American Medical Informatics Association, 31:583–590, 2024. doi: 10.1093/jamia/ocad247

[19] P. Ryan. Our journey. where the ohdsi community has been and where we are going, 2025.

[20] G. Schwarzer. meta: An r package for meta-analysis. R News, 7(3):40–45, 2007.

[21] B. Shneiderman. The eyes have it: A task by data type taxonomy for information visualizations. In Proc. IEEE Symposium on Visual Languages, pp. 336–343, 1996. doi: 10.1109/VL.1996.545307

[22] SNOMED International. Snomed ct technical implementation guide, 2024.

[23] E. A. Voss, A. Shoaibi, Y. S. Lai, C. Blacketer, T. Alshammari, K. Berencsi et al. Contextualising adverse events of special interest to characterise the baseline incidence rates in 24 million patients across 26 databases. eClinicalMedicine (Lancet), 58:101932, 2023. doi: 10.1016/j.eclinm.2023.101932

[24] J. Walonoski, M. Kramer, J. Nichols, A. Quina, C. Moesel, D. Hall et al. Synthea: An approach, method, and software mechanism for generating synthetic patients and the synthetic electronic health care record. Journal of the American Medical Informatics Association, 25(3):230–238, 2018. doi: 10.1093/jamia/ocx079

[25] Q. Wang and R. S. Laramee. Ehr star: The state-of-the-art in interactive ehr visualization. Computer Graphics Forum (EuroVis STAR), 41(1):69–105, 2022. doi: 10.1111/cgf.14424

[26] L. Wilkinson and M. Friendly. The history of the cluster heat map. The American Statistician, 63(2):179–184, 2009. doi: 10.1198/tas.2009.0033

[27] World Health Organization. International classification of diseases, 10th revision (icd-10), 2019.

[28] World Health Organization. Anatomical therapeutic chemical (atc) classification, 2024.

